# Using A Socio-Ecological System (SES) Framework to Explain Factors Influencing Countries’ Success Level in Curbing COVID-19

**DOI:** 10.1101/2020.11.17.20226407

**Authors:** Nur Amiera Suhud, Gabriel Hoh Teck Ling, Pau Chung Leng, AK Muhamad Rafiq AK Matusin

## Abstract

Little attention has been paid to interactions between institutional-human-environment dimensions, which are believed to impact the outcome of COVID-19 abatement. Thus, through the diagnostic SES framework analysis, this paper aims to investigate what and how the multifaceted social, physical, and governance factors affect the success level of 7 selected Asia-Pacific countries (namely South Korea, Japan, Malaysia, Singapore, Vietnam, Indonesia, and New Zealand) in combatting the COVID-19 pandemic. Drawing on secondary data from February 2020 to June 2020, the success or severity level of a country was measured by cumulative positive cases, average daily increase, and the mortality rate. A qualitative content analysis, covering code assignation, i.e., Present (P), Partially Present (PP), Absent (A) for each SES attribute, as well as rank ordering (from 1^st^ to 7^th^) and score calculation (from 3 to 21) for the success level between the countries, was undertaken. Attributes (design principles) of SES factors, such as past experiences facing similar diseases, facilities mobility, lockdown measures, penalty, and standard of procedures in public spaces are deemed significant in determining the abatement outcome or severity of a country. The findings show that Vietnam (1^st^) and New Zealand (2^nd^) adopting most of the design principles of governance (with the scores of 15 and above) had successfully eliminated the virus, while Indonesia (7^th^) and Japan (6^th^) were deemed least successful (scoring between 3-9), likely due to the low presence frequency of design principles. Not only does the study validate SES framework adaptability in a health-related (non-commons) setting, where some design principles used in resource/commons governance are also relevant in explaining the COVID-19 outcome, the critical attributes of institutional-social-ecological factors are highlighted, ultimately helping policymakers devise more strategic measures to address the crisis.

## INTRODUCTION

COVID-19 was unknown to the masses prior to its first case in December 2019 (World Health Organization, 2020b) although there were speculations that the earliest case dated back to November 2019 (Brynner, 2020; Li et al., 2020). It was first regarded as viral pneumonia but was then analysed to be a viral infection and has the capability to be transmitted through human to human interaction (Shereen et al., 2020).

As of June 2020, there were countries that had successfully mitigated the problem, while some were struggling and had failed due to countless reasons. Much emphasis has been put on the science and pharmaceutical aspect in effort to produce vaccines to cure the pandemic, although it is believed that social behaviours, despite being intangible, indirect and qualitative, leaning towards prosociality, via a cooperative action can also contribute to curbing the pandemic (Ling & Ho, 2020). Through this research, an integration between health and the systemic institutional-social-ecological (SES) framework was carried out, apart from knowing its relevancy when applied outside its commons domain, specifically to identify answers to the following research question— How the diagnostic IAD-SES^1^ framework can help answer the level of success or failure of countries in curbing the pandemic? It has come to attention that there is a noticeable gap of research in exploring what and how institutional-social-ecological dimensions of the Institutional Analysis and Development (IAD) or SES framework help understand the unprecedented crisis. More precisely, given with numerous exogenous factors, this study identifies which SES attributes (design principles) are significant and effective in explaining and thus curbing the pandemic. The framework, synonymous with commons and resource management or collective action, is rarely applied in a health and disease-related topic, such as COVID-19; relationships between the transmission of COVID-19, socio-economic and environmental factors should be demystified in order to devise a holistic strategy in mitigating the problem.

The Institutional Analysis and Development (IAD) and SES framework are both founded by Elinor Ostrom (1990). The former, mostly adopted by social scientists, is primarily used to evaluate the effects of alternative institutional arrangements. It was envisioned as a tool for scholars of different disciples to communicate with each other regardless of their broad perspective to pave a way towards better understanding of a situation (Cole et al., 2019). Within the framework, it is important to understand the action situation/interaction that actors are in, plausible choices made by them and how it will affect the pattern of the outcome. In order to predict choices that will be made, it is crucial to know: (1) the resources brought into the situation, (2) the valuation assigned by the actor to the state of the world and to the action; (3) how the actors acquire, process, retain and use the knowledge and information; and (4) ways used by the actor in selecting a particular decision (Mcginnis et al., 1992).

The IAD framework, however, is criticized as it lacks in terms of diversity and complexity of natural system and processes (Cole et al., 2019). Therefore, SES is an improved version building upon the IAD framework by expanding the basic variables into more relevant categories as shown in Figure 1. The SES framework provides a more detailed variable oriented analysis of the social-ecological system (Cole et al., 2019). The community and governance attributes in the IAD context have been maintained as social and governance systems in an SES, while biophysical/ecological attributes converge into two sections, which are resource systems and resource units. SES, normally applied in the context of commons or CPRs governance, is proven to be versatile and adaptable, due to its generality. It also enables a comparison of different settings in a study where data are collected with the means to compare (Partelow, 2018).

**FIGURE 1.**
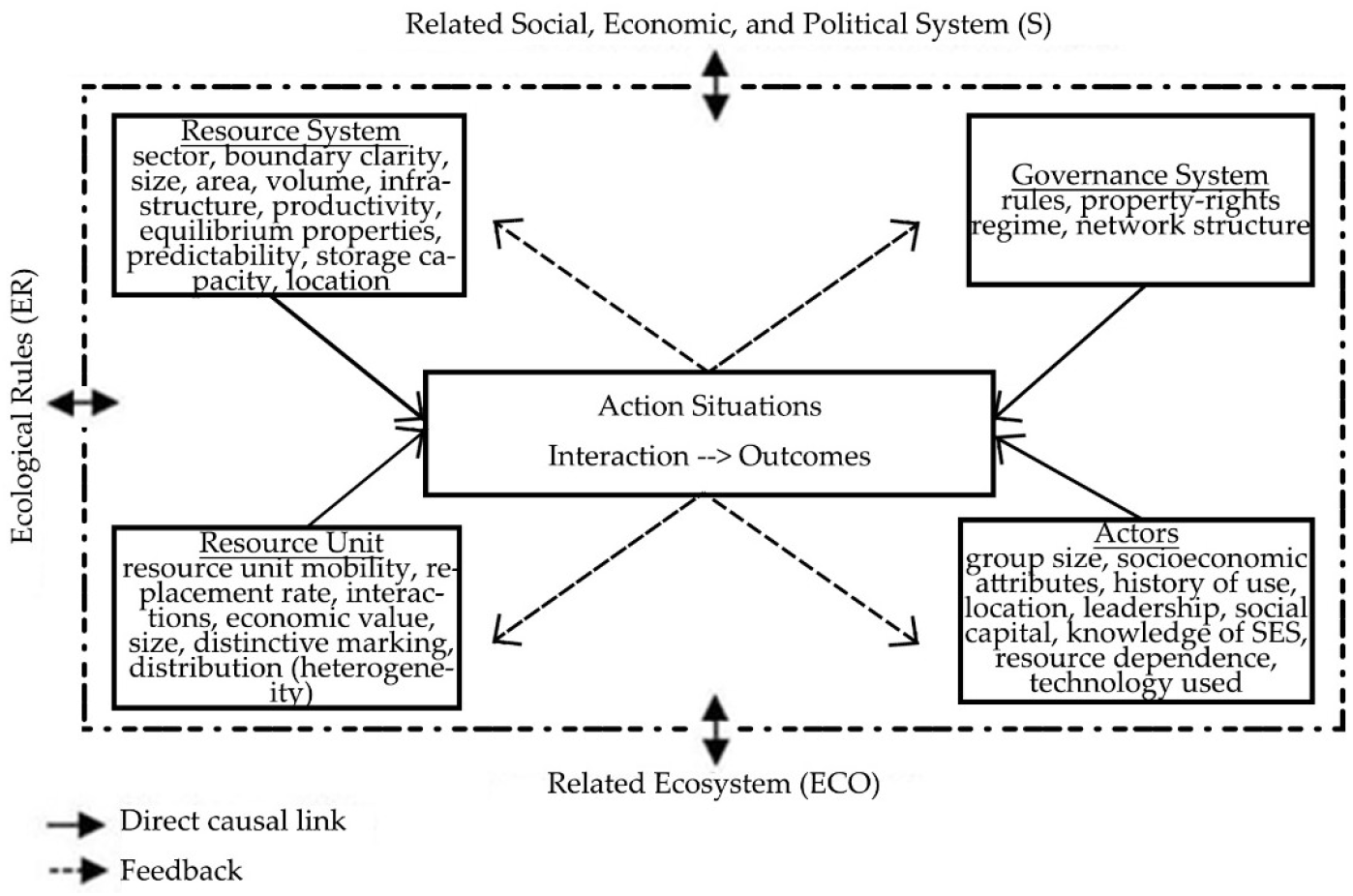
Second and third tier SES attributes. Adapted from Ostrom and Cox (2010)

By adopting the SES framework, it allows researchers to have an organized variable-oriented and process-oriented line of arguments, involving systematic networks of action situations, and hence a more informed decision (Cole et al., 2019; Tashman, 2020). When adapted into the context of COVID-19, the institutional-socio-ecological attributes (e.g., penalty, lockdown, facilities, technology, and population density, and past knowledge) and their interaction (e.g., monitoring, communication, swab testing, containment enforcement) would influence the number of cumulative cases, average daily increase, and the mortality rate and thus translate the success level of a country into low, medium and high, as diagrammatized in Figure 2.

**FIGURE 2.**
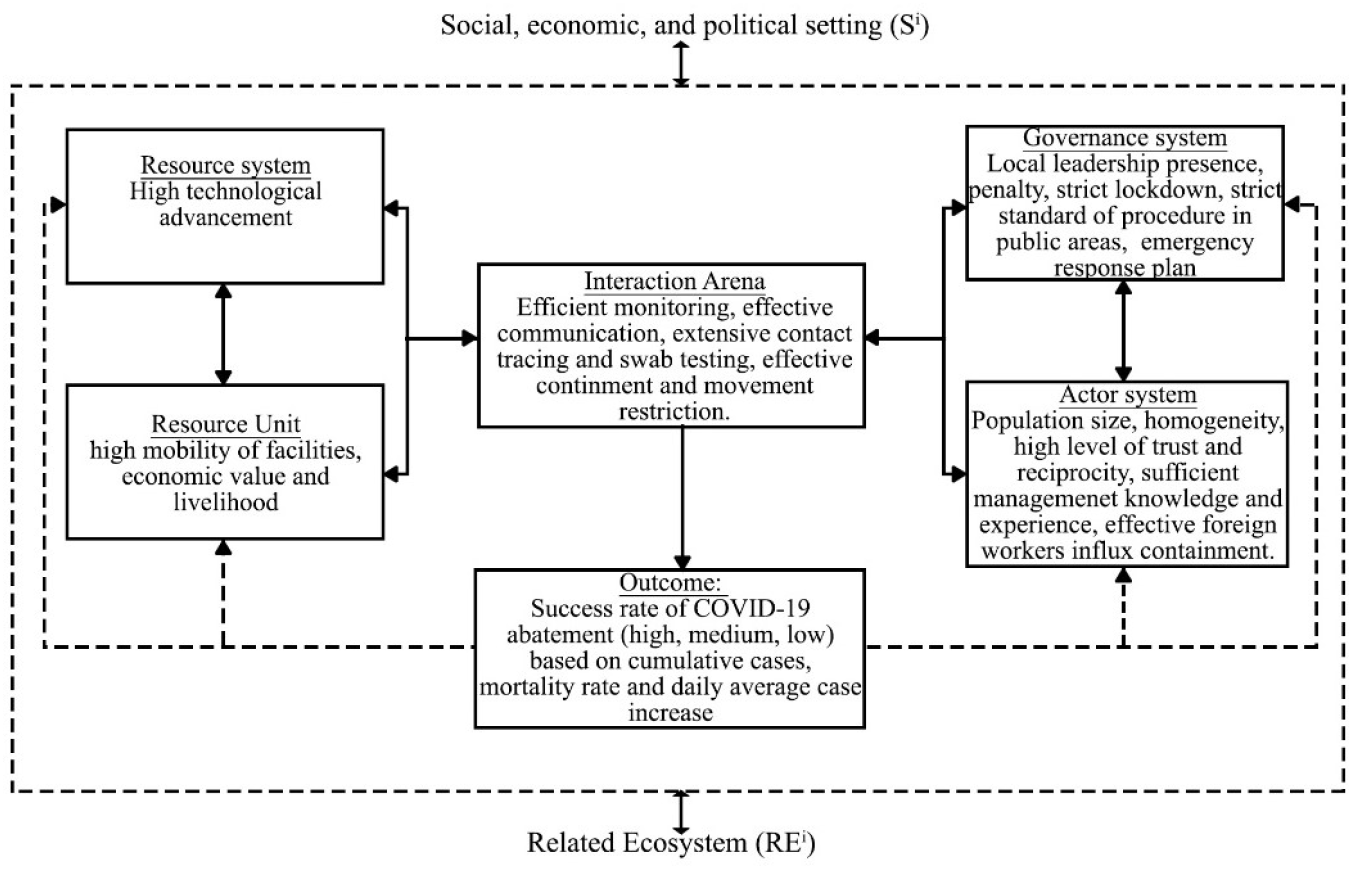
The SES framework in the COVID-19 context. Source: Adapted from McGinnis and Ostrom (2014)

Theoretically, design principles (DP) or critical success factors that are present in a successful resource management should also exist in successful pandemic containment and mitigation. The hypothesis was that the higher the amount/frequency of design principles (successful attribute) that are present or occur in countries, the higher the success level of them curbing the pandemic. Despite all these, research focusing on socio-economic and socio-ecological drivers of the COVID-19 transmission remains scarce when in fact, it does hold an equally important role as a critical determinants of COVID-19 transmission risks. Although few studies link both socio-ecological and climate factors with transmission levels of COVID-19, a study had proven that the latter (climatic factor) does not affect significantly, and hence its absence in this paper (Su et al., 2020). Therefore, this study attempts to contribute theoretically and methodologically by expanding the application of the SES framework and its institutional-social-ecological design principles in the COVID-19 context. Studying these factors should also be of practical significance because the findings can serve as a guide/reference that helps forecast the COVID-19 outcome more accurately as to whether a country is likely to fail or succeed in addressing the pandemic. Ultimately, this study enables researchers and policymakers to devise more holistic and effective strategies to reduce and prevent similar incidents in the future. The following sections of the paper further elaborates the topic in terms of (i) study areas, (ii) methods of data collection and analysis, (iii) results, findings, and discussions, and (iv) lastly a conclusion.

## METHODOLOGY

### STUDY AREAS

Via a thorough literature review a comparative study was carried out between the selected countries located in Asia Pacific. This study has enlisted Japan, Korea, Malaysia, Singapore, Vietnam, Indonesia, and New Zealand as study areas and classified them into three categories, namely countries (i) that had successfully flattened the curve, (ii) attempting to flatten the curve, and (iii) that failed to flatten the curve. Such categorization was based on their cumulative cases, daily average increase and mortality rate. Table 1 illustrates an overall data of the three criteria for each country. In general, Vietnam fared the best with 0.9 average cases as of 30^th^ June and 0% mortality rate as illustrated in Figure 4 and Figure 5. Figure 3 has shown that the cumulative cases of Vietnam were also at the lowest, i.e., 355. The government’s swift and strict measures had proven to be successful as the country was one of the earliest to eliminate the virus (Roser et al., 2020). Meanwhile, Indonesia recorded a cumulative case of 49 000 with a daily average increase of 1000 cases, and these number was the highest among the seven countries. However, its mortality rate comes second after Japan. Next paragraph provides brief background for each country as to how COVID-19 cases started to emerge.

**TABLE 1:**
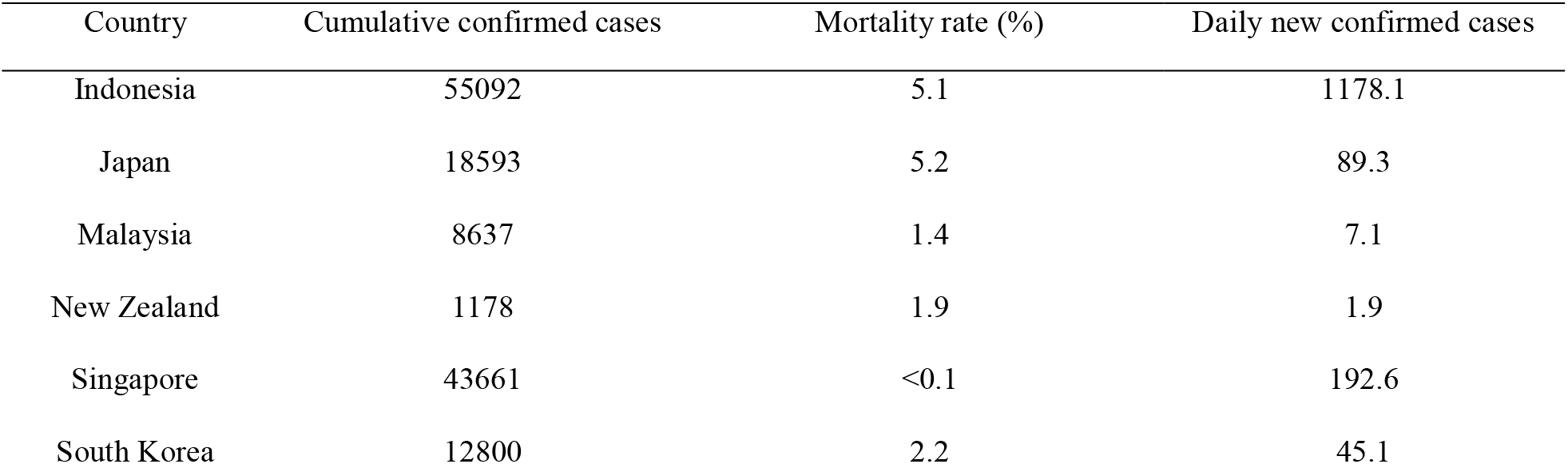
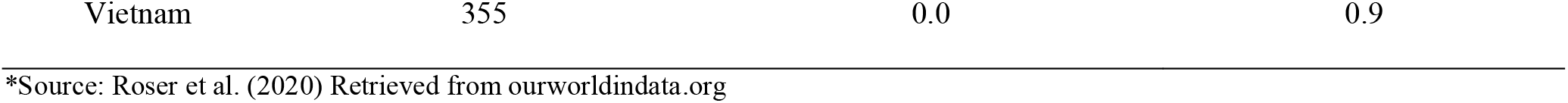
Data of cumulative cases, daily average increase, and the mortality rate as of 30^th^ June 2020.

**FIGURE 3:**
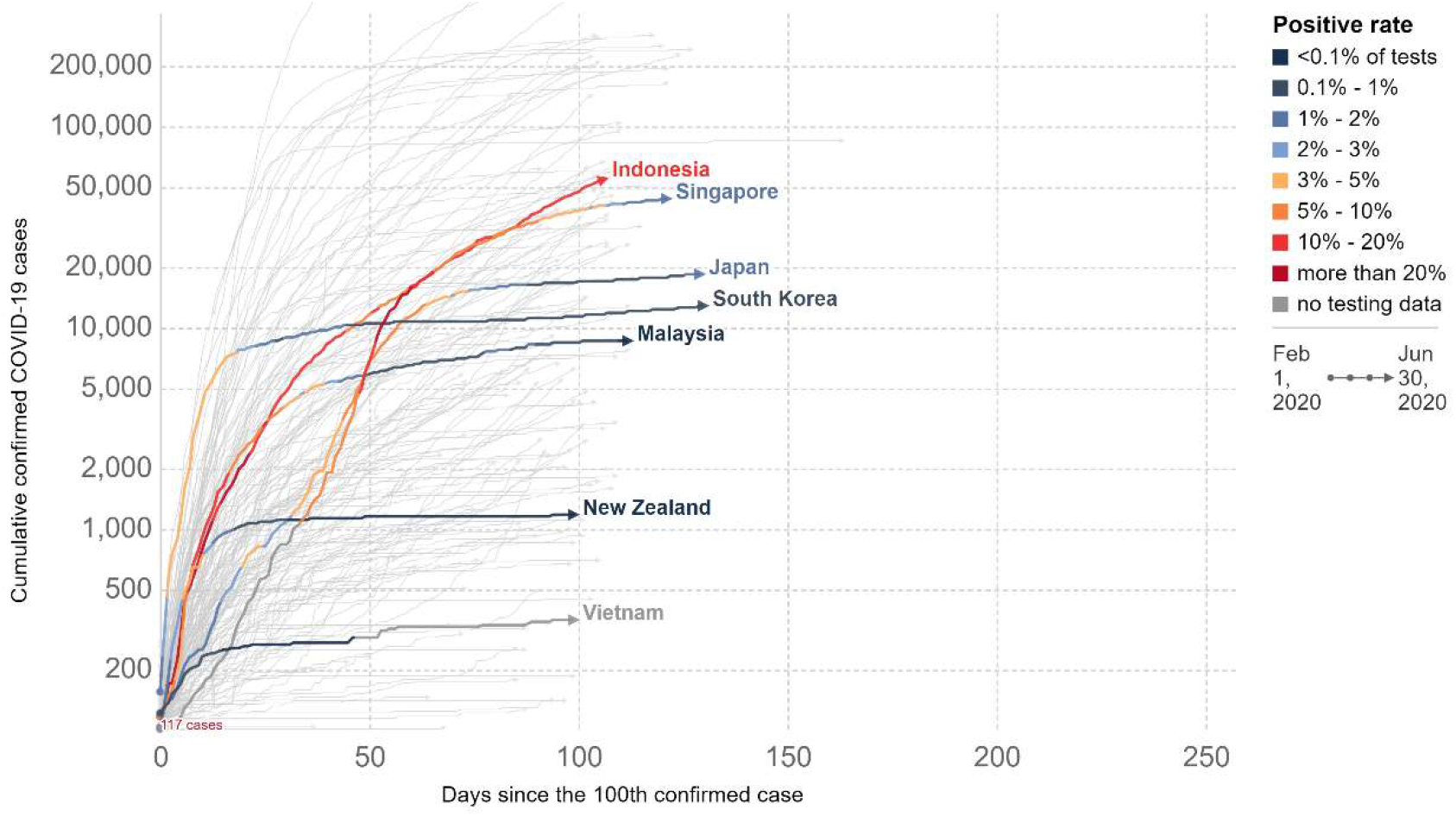
Cumulative COVID-19 cases. Source: Roser et al. (2020) Retrieved from ourworldindata.com

**FIGURE 4:**
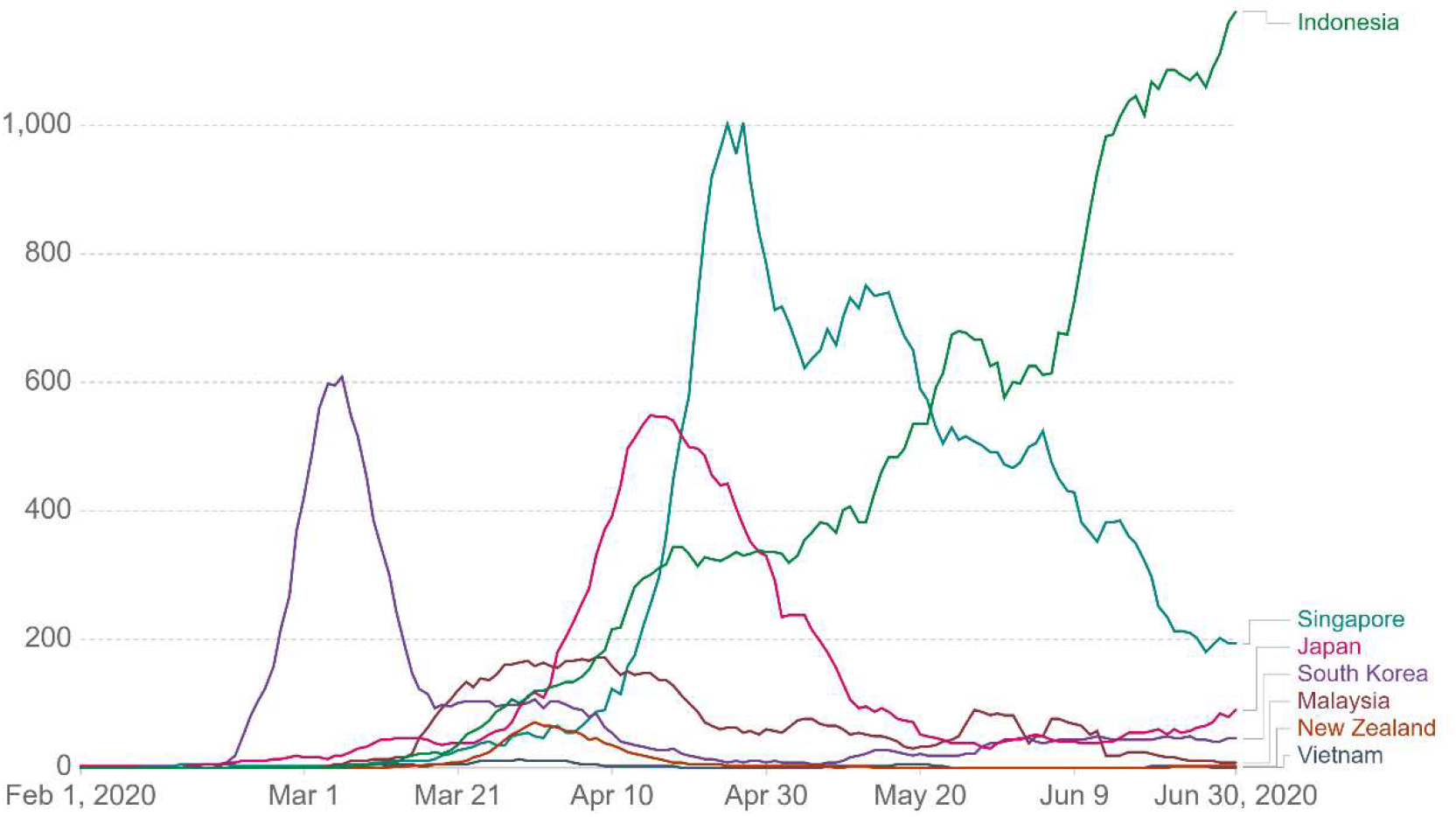
Daily average case increase (7 days average). Source: Roser et al. (2020) Retrieved from ourworldindata.com

**FIGURE 5:**
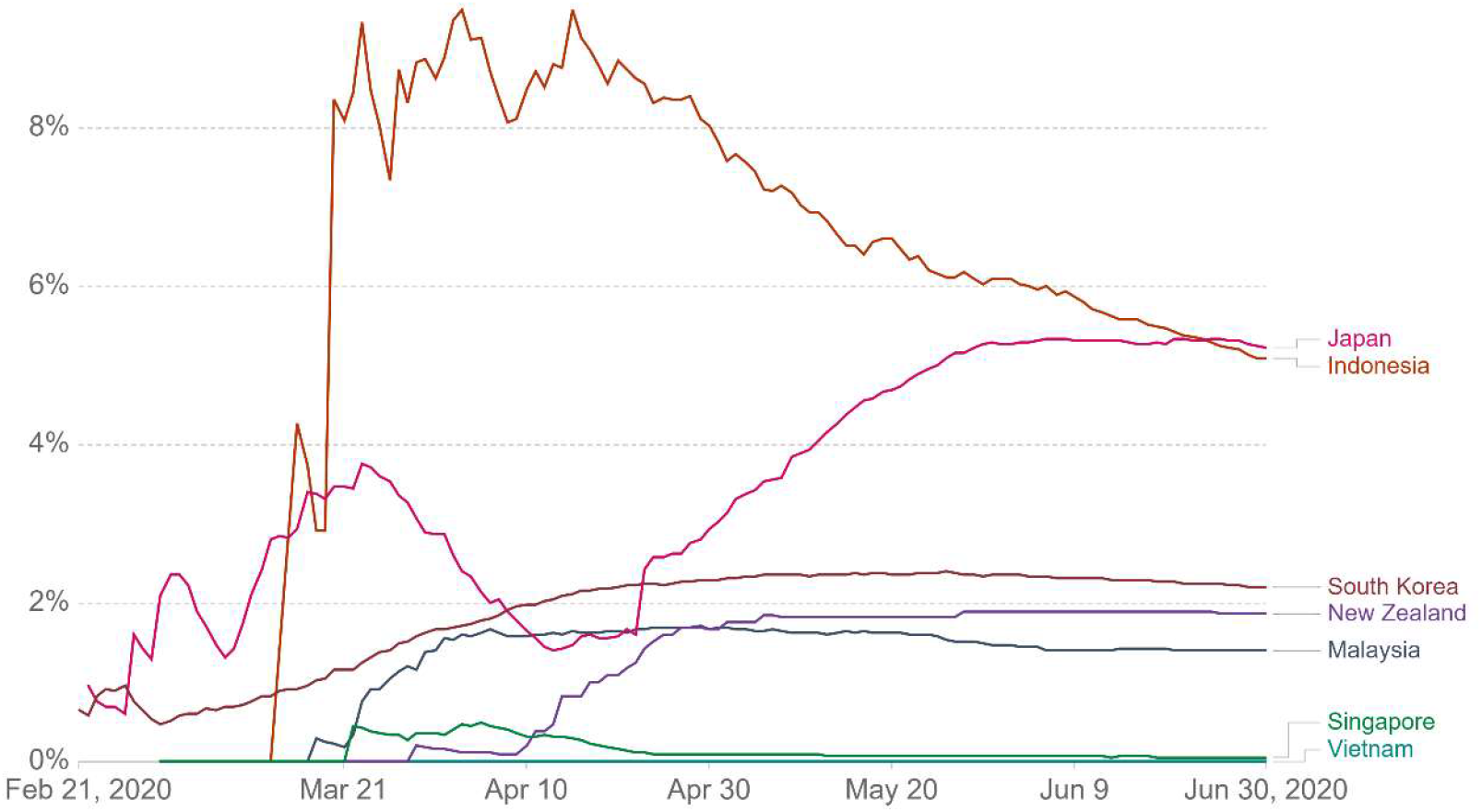
Mortality rate of each country. Source: Roser et al. (2020) Retrieved from ourworldindata.org

Cases of COVID-19 had first emerged in these countries from early January to the end of February. In some countries, COVID-19 was brought in by Chinese nationalities or individuals returning from China, such as in Singapore, New Zealand, and Japan (Henrickson, 2020; World Health Organization, 2020a). Indonesia recorded its first case after a nationality came in contact with an infected person in Malaysia (Channel News Asia, 2020b). Even so, Japan was the first to record a human-to-human transmission as its first case did not visit the market but was in close contact with a pneumonia patient (World Health Organization, 2020a). In some countries, the first case did not necessarily lead to a booming number of cases. In Malaysia, cases started to skyrocket due to a religious event in Sri Petaling in March, attended by almost 16000 individuals from Malaysia and others (Free Malaysia Today, 2020). The same phenomenon occurred in South Korea when an elderly woman in Daegu was tested positive for the virus. She attended a 10000 member mass gathering at the Shincheonji Church (Levkowitz, 2020).

### DATA COLLECTION

To execute a comparative study, we had outlined at total of 14 attributes, such as population size and density, homogeneity, trust and reciprocity, local management knowledge and experience, foreign worker clusters influx containment, mobility, technology, economy, local leadership, penalty, lockdown, standard of procedure (SOP) in public areas, and emergency response plan. These attributes were based on the IAD-SES framework which had been localized to the COVID-19 context. This study made use of secondary sources, such as research papers, government documents, newsletters, and official websites. Keywords such as ‘IAD-SES framework’, ‘Social-ecological system’, ‘COVID-19’ were used on Google Scholar to garner information. Other than that, we also referred to ourworldindata.org to collect statistical data on cumulative cases, mortality rate, testing rates, and daily average increase of cases. The Government Response Stringency Index was also referred to as it was used as important data input for the “interaction or action arena” of the SES framework, prior to explaining the COVID-19 outcome of a country. It is worth noting that data extracted only cover information from the month of February 2020 to June 2020. The specific timeline was chosen as most country cases were at a peak during that period.

### DATA ANALYSIS

The study employed a qualitative content analysis to study institutional-social-ecological attributes, using Ostrom’s 8 design principles (Ostrom, 1990) as one of the theoretical underpinnings. To aid the analysis, a coding system was used to determine IAD based SES attributes in each country. According to suitability and relevancy, each attribute was assigned with a code based on a ratio or percentage of DP presence/occurrence, i.e., Absent (A) (with 0%-29%), Partially Present (PP) (with 30%-69%) or Present (P) (with 70%-100%). The coding process, adapted from Gari’s et al., (2017) methodology, was undertaken in a rather intuitive or subjective manner, especially when data were inconsistent and unavailable. Ultimately, in line with Ostrom’s successful commons governance focusing on the importance of the presence of design principles, a higher frequency/occurrence of P indicates a better COVID-19 containment, and therefore a higher success rate of the country, while a lower count of P connoting a higher frequency of A or PP means otherwise.

At the same time, the success level of a country was measured and determined based on the total scores obtained from the three criteria, namely the cumulative number of cases of each country, the day to day average increase, and the mortality rate of the country. More precisely, the total scores were calculated based on the ranking (i.e., 1^st^ to 7^th^) and values assigned (from 1 to 7) to each criterion. Table 2 summarizes the options of values for each rank as well as the score ranges (from 3 to 21) for the high, medium, and low outcomes, respectively.

**TABLE 2:** Values for each rank and the range of score determining the outcome

| Rank | Value | Score | Outcome |
| --- | --- | --- | --- |
| 1<sup>st</sup> | 7 | 16-21 | High |
| 2<sup>nd</sup> | 6 | 10-15 | Medium |
| 3<sup>rd</sup> | 5 | 3-9 | Low |
| 4<sup>th</sup> | 4 |  |  |
| 5<sup>th</sup> | 3 |  |  |
| 6<sup>th</sup> | 2 |  |  |
| 7<sup>th</sup> | 1 |  |  |

## RESULTS AND DISCUSSIONS

Based on the analysis, a summarized result spanning 7 countries with respective SES attributes is shown in Table 3. Detailed result explanations are provided below, based on each SES attribute.

**TABLE 3.** Summarized institutional-socio-ecological attributes for seven countries.

| IAD- based SES Attributes | New Zealand | Singapore | Vietnam | Malaysia | South Korea | Japan | Indonesia |
| --- | --- | --- | --- | --- | --- | --- | --- |
| Community/social attributes |  |  |  |  |  |  |  |
| Population size and density | 4.886 million (2020)<br>18 per km<sup>2</sup> (2020) | 5.6 million (2020)<br>8358 per km<sup>2</sup> (2020) | 97 million (2020)<br>314 per km<sup>2</sup> (2020) | 32.5 million (2020)<br>99 per km<sup>2</sup> (2020) | 51 million (2020)<br>527 per km<sup>2</sup> (2020) | 126.5 million (2020)<br>347 per km<sup>2</sup> (2020) | 274.2 million (2020)<br>151 per km<sup>2</sup> (2020) |
| High homogeneity | PP | PP | P | PP | P | P | PP |
| High level of trust and reciprocity | P | P | P | PP | PP | A | PP |
| Sufficient local management knowledge and experience | P | P | P | P | P | P | PP |
| Effective foreign workers influx containment | N/A | PP | N/A | A | N/A | N/A | N/A |
| Spatial/physical system attributes |  |  |  |  |  |  |  |
| High mobility | PP | P | PP | PP | P | PP | A |
| High technological advancement | P | P | P | PP | P | PP | PP |
| Economic/GDP/GNI (livelihood) | High Income<br>GDP: 52<br>GNI: 30 (purchasing power parity)<br>24 (atlas methodology)<br>15 percent lived in poverty in 2010.<br>(Belammy, 2011) | High Income<br>GDP: 34<br>GNI: 3 (purchasing power parity)<br>11 (atlas methodology)<br>Based on research, \$1878 for absolute poverty and \$5076 for relative poverty threshold | Lower mid-income<br>GDP: 45<br>GNI: 127 (purchasing power parity)<br>141 (atlas methodology)<br>6.7% living under national poverty line. (2018) | Upper mid income<br>GDP: 36<br>GNI: 53 (purchasing power parity)<br>68 (atlas methodology)<br>Changes from RM980 to RM2208.5. 6% for absolute poverty | High Income<br>GDP: 12<br>GNI: 29 (purchasing power parity)<br>30 (atlas methodology)<br>Almost 15%. Prominent in old people. (2012) | High Income<br>GDP: 3<br>GNI: 26 (purchasing power parity)<br>27 (atlas methodology)<br>15.7% o with household income less than half of median of the entire population. (2020) | Upper mid income<br>GDP: 16<br>GNI: 109 (purchasing power parity)<br>118 (atlas methodology)<br>9.6% / 25 million live below the national poverty live and 40% (180 million) living just above that line (2020) |
| Governance/institution attributes |  |  |  |  |  |  |  |
| Local leadership presence | P | PP | P | PP | PP | PP | PP |
| Penalty | P | P | P | P | P | A | P |
| Strict lockdown | P | P | P | P | PP | A | PP |
| Strict standard of procedure in public areas | P | P | P | PP | P | P | P |
| Emergency response plan | P | P | P | P | P | P | P |
| Interaction Arena |  |  |  |  |  |  |  |
| Efficient monitoring | P | P | P | P | P | A | P |
| Effective communication | P | P | P | P | P | P | P |
| Extensive contact tracing and swab testing (per thousand) | 1.31 | 1.92 | 1.31 | 0.32 | 0.22 | 0.06 | 0.04 |
| Effective containment and movement restriction | P | P | P | P | P | A | P |
| Outcome | CC: 1178 (2<sup>nd</sup>)<br>DAI: 1.9 (2<sup>nd</sup>)<br>MR: 1.9% (4<sup>th</sup>) | CC: 43661 (6<sup>th</sup>)<br>DAI: 192.6 (6<sup>th</sup>)<br>MR: <0.1% (2<sup>nd</sup>) | CC: 355 (1<sup>st</sup>)<br>DAI: 0.9 (1<sup>st</sup>)<br>MR: 0.0% (1<sup>st</sup>) | CC: 8637 (3<sup>rd</sup>)<br>DAI: 7.1 (3<sup>rd</sup>)<br>MR: 1.4% (3<sup>rd</sup>) | CC: 12800 (4<sup>th</sup>)<br>DAI: 45.1 (4<sup>th</sup>)<br>MR: 2.2% (5<sup>th</sup>) | CC: 18593 (5<sup>th</sup>)<br>DAI: 89.3 (5<sup>th</sup>)<br>MR: 5.2% (7<sup>th</sup>) | CC: 55092 (7<sup>th</sup>)<br>DAI: 1178.1 (7<sup>th</sup>)<br>MR: 5.1% (6<sup>th</sup>) |
| Score | 16 | 10 | 21 | 15 | 11 | 7 | 4 |
| Success level of COVID-19 abatement | High | Medium | High | Medium | Medium | Low | Low |

CC: Cumulative cases
DAI: Daily average increase of cases
MR: Mortality rate

### POPULATION SIZE AND DENSITY

Based on a study done by Kadi and Khelfaoui in Algeria, population density indeed plays a role in the transmission of virus (Kadi & Khelfaoui, 2020). However, it cannot be a strong explanatory factor as to why the number of cases are as known (Bouba-Olga, 2020). Among the 7 countries, Singapore has the highest population density (8358 per km^2^) as shown in Table 3. Despite the high population density, the country having adequate facilities and economic strength managed to keep the number of positive cases and the death rate under control. In contrast, Indonesia fumbled despite having a lower population density due to the shortage of facilities and economy.

### HIGH SOCIAL HOMOGENEITY

Based on Table 3 above, Vietnam, South Korea, and Japan were justified as P because their populations appear to be relatively homogenous in terms of culture and some demographic attributes (ethnics and language). Meanwhile, other countries were considered as PP due to their multicultural and racial settings, such as Malaysia, Indonesia, New Zealand, and Singapore. Culturally, Japanese had always upheld a high standard of hygiene and obedience. They have always practiced social distancing as they respect personal space. Wearing masks when feeling sick is a norm to prevent contracting the virus to others (Tashiro & Shaw, 2020). Therefore, there are no objections among its citizens when mask rules were enforced by the government. However, to posit the homogeneity of a nation contributes positively to the rate of success, it seems rather farfetched as the analysis had shown that it does not play a major role.

### HIGH LEVEL OF TRUST AND RECIPROCITY

Table 4 is an extension of Table 3, explaining the level of trust and reciprocity owned by each country. Having trust from the people ensures that the public follows the rules undisputedly (i.e., higher compliance), allowing the leaders to convince them of mass testing and quarantines before it gets worse and hence keeping the virus at bay earlier on (Kleinfeld, 2020). In other words, having citizens’ high trust towards the government may contribute positively to the success level of a country in curbing the pandemic. The trust level of each country was based on Edelman’s Barometer report 2020 and the Guardian. For example, countries like Singapore and New Zealand are deemed to have trust due to transparency and the leader’s excellent risk communication. Meanwhile, other countries’ governments (Malaysia, South Korea and Vietnam) had constantly proven their competency, hence garnering public trust in their capabilities (Hong, 2020; Kleinfeld, 2020; Levkowitz, 2020; Tran et al., 2020; Woo, 2020). In Vietnam, the people believe that the government is working for the betterment of the country; the Vietnamese government is also competent in providing prompt communication and medical supplies thus allowing them to carry out strategies similar to wealthier countries despite being a lower income nation (Kleinfeld, 2020).

**TABLE 4.** Citizen’s trust towards the government handing the pandemic.

|  | Indonesia | Japan | Malaysia | New Zealand | Singapore | South Korea | Vietnam |
| --- | --- | --- | --- | --- | --- | --- | --- |
| Trust Level | Most trust | Distrustful | Neutral | Most trust | Most trust | Neutral | N/A |

\*Source: (Edelman, 2020; The Guardian, 2020)

Despite being placed in the most trust category (Table 4), the Republic of Indonesia has been speculated to hide the real number of cases in the country by activists and political opponents (Aljazeera, 2020). This had led us to code Indonesia PP for this attribute. Japan’s government too, faced the same problem. The government was labelled distrustful by the barometer, and this was further backed up by a poll done by NHK, where the survey stated that more than half of the population disapproved the Prime Minister’s and his government’s ways in handling the pandemic (The Economist, 2020).

### SUFFICIENT LOCAL MANAGEMENT KNOWLEDGE AND EXPERIENCE

Sufficient local management knowledge and experience are mostly present in all seven countries as seen in Table 3. It was found that most countries have had experiences in handling outbreaks, such as SARS, Measles and H1N1 (Table 5). These experiences helped them improve their healthcare system in infection prevention and control, activate response protocols, increase number of thermal scanners at all borders and realized the importance of isolating infected cases and quarantine measures (Bernama, 2020; Hong, 2020; Rahman, 2020). This means that countries with the past experiences in handing related pandemic contagions can better cope with COVID-19. Contrary to other countries, Japan had opted for lesser testing based on its experience in handling a pandemic. Previous outbreaks had seen the public flocking the hospitals to get tested increasing the transmission risks. Thus, the Ministry of Health, Labour and Welfare was concerned that crowded hospital lobbies would lead to a surge of COVID-19 cases and hence finalized their decision to limit accessibility (Nippon, 2020).

**TABLE 5.** Countries’ past experiences in handling pandemics.

|  | Indonesia | Japan | Malaysia | New Zealand | Singapore | South Korea | Vietnam |
| --- | --- | --- | --- | --- | --- | --- | --- |
| SARS | Yes | Yes | Yes | Yes | Yes | Yes | Yes |
| MERS | No | No | Yes | No | No | Yes | No |
| Nipah Virus | Yes | No | Yes | No | Yes | No | No |
| H1N1 | Yes | Yes | Yes | Yes | Yes | Yes | Yes |
| Measles | Yes | Yes | Yes | Yes | Yes | Yes | Yes |
| 1918 flu pandemic | Yes | Yes | Yes | Yes | Yes | Yes | No |

\*Source: (Levkowitz, 2020; Rahman, 2020; The Encyclopedia of New Zealand, 2020; Woo, 2020; World Health Organization, 2018a)

### EFFECTIVE FOREIGN WORKERS INFLUX CONTAINMENT

Table 3 had stated that both Malaysia and Singapore had a large influx of foreign workers which leads to clusters of COVID-19 cases. If there is a proper containment system of the worker, it would be indicated as P. On the other hand, other countries marked N/A which means this attribute is not applicable as they did not have a huge influx of foreign workers leading to COVID cluster emergence. A proper containment system helps eradicate possibilities of a surge of cases therefore controlling the number of cumulative cases and maintaining a low mortality rate.

The rise of a foreign worker cluster in Singapore was apparent from 15^th^ to 30^th^ March, with almost 61% imported cases. Singapore has over a million foreign workers with most of them living in cramped housing (Koh, 2020). Both Malaysia and Singapore had cited poor living condition as a factor that had sparked a widespread infection among the group (Malay Mail, 2020b). The severity of cases in Malaysia had forced the government to employ a more stringent lockdown, called the Enhanced Movement Control Order (EMCO) (Sandanasamy et al., 2020).

### HIGH FACILITIES MOBILITY

In determining the code frequency in Table 3, P translates to countries with a high healthcare facility mobility, while PP translates to a moderate mobility, and A translates to a low facilities mobility. Having an ample amount of resources (physicians and facilities) allows more patients to be treated at one time, hence ensuring the best healthcare services to all. Therefore, the COVID-19 mortality rate could be lowered. Generally, high-income countries like South Korea, Singapore and Japan possessed sufficient health facilities and manpower (Table 6), except for New Zealand and Japan (Mazey & Richardson, 2020; The Conversation, 2020). However, a lower income country like Vietnam has managed to control the pandemic due to its governments’ swift strategy as they recognized its inability to contain a high amount of cases if the pandemic get serious (Jones, 2020).

**TABLE 6.**
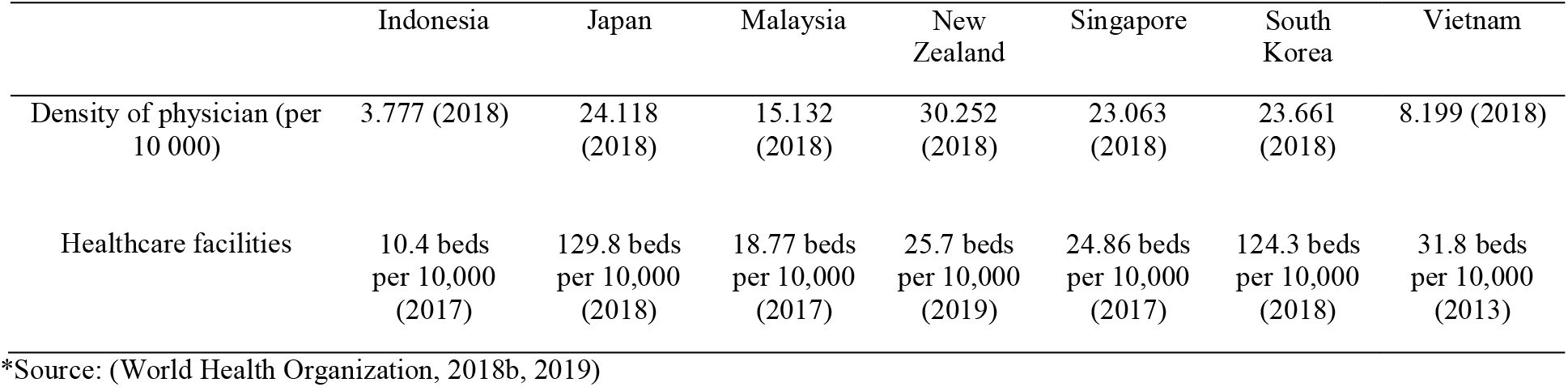
Components that affects the mobility of facilities.

|  | Indonesia | Japan | Malaysia | New Zealand | Singapore | South Korea | Vietnam |
| --- | --- | --- | --- | --- | --- | --- | --- |
| Density of physician (per 10 000) | 3.777 (2018) | 24.118 (2018) | 15.132 (2018) | 30.252 (2018) | 23.063 (2018) | 23.661 (2018) | 8.199 (2018) |
| Healthcare facilities | 10.4 beds per 10,000 (2017) | 129.8 beds per 10,000 (2018) | 18.77 beds per 10,000 (2017) | 25.7 beds per 10,000 (2019) | 24.86 beds per 10,000 (2017) | 124.3 beds per 10,000 (2018) | 31.8 beds per 10,000 (2013) |

\*Source: (World Health Organization, 2018b, 2019)

Indonesia has the lowest density/number of physicians among the seven countries. In addition, its facilities are in a dire condition (Pisu, 2010). The country’s Ministry of Health had recognized an uneven distribution of health facilities and quality across Indonesia (Pisu, 2010; Setiati & Azwar, 2020). Some sub-districts in Indonesia did not possess any health centre and lack necessities such as electricity, clean water, and proper equipment with limited transportation (United Nations Office for the Coordination of Humanitarian Affairs & The United Nations Resident Coordinator Office, 2020). Obvious disparity between rural areas and urban regions is common in many countries i.e., Kuala Lumpur. Facilities in these areas are usually government funded with fewer doctors hence a lower doctor-to-patient ratio (Falcon, 2019).

### HIGH TECHNOLOGICAL AVAILABILITY

The presence of technology aids governments and healthcare sectors devise strategies towards the abatement of the virus. In Table 3, countries with code P has a comprehensive contact tracing, medical equipment productions as well as developing its own test kits. Having PP for this attribute means limited tracing or the absence of medical equipment and test kit production. Seen in Table 7, most of the countries had opted for comprehensive tracing, while Indonesia’s and Japan’s testing capacity was limited (Roser et al., 2020). A comprehensive tracing allows more cases to be detected (or swab tested) which can be done through integration of an application. The efficiency of contact tracing applications is high as a study shows that MySejahtera has uncovered 251 confirmed COVID-19 cases or 3% of 8308 cases as of June (Boo, 2020).

**TABLE 7.**
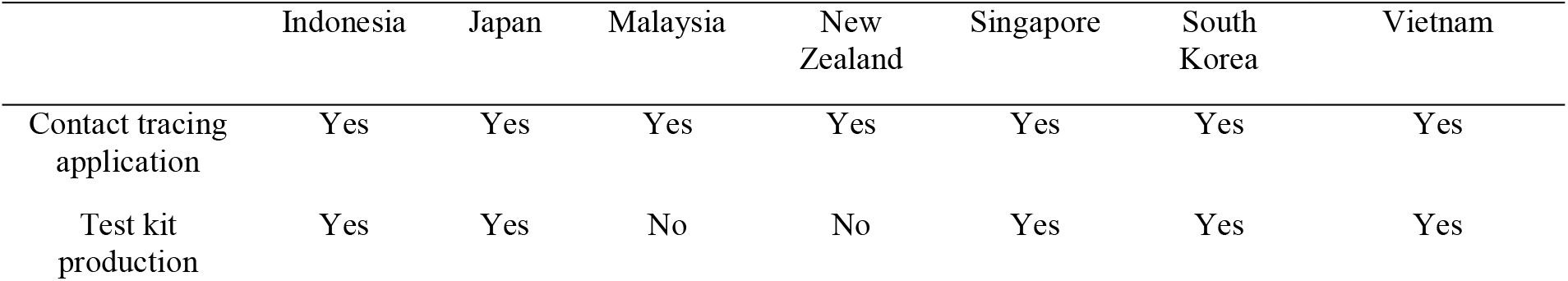
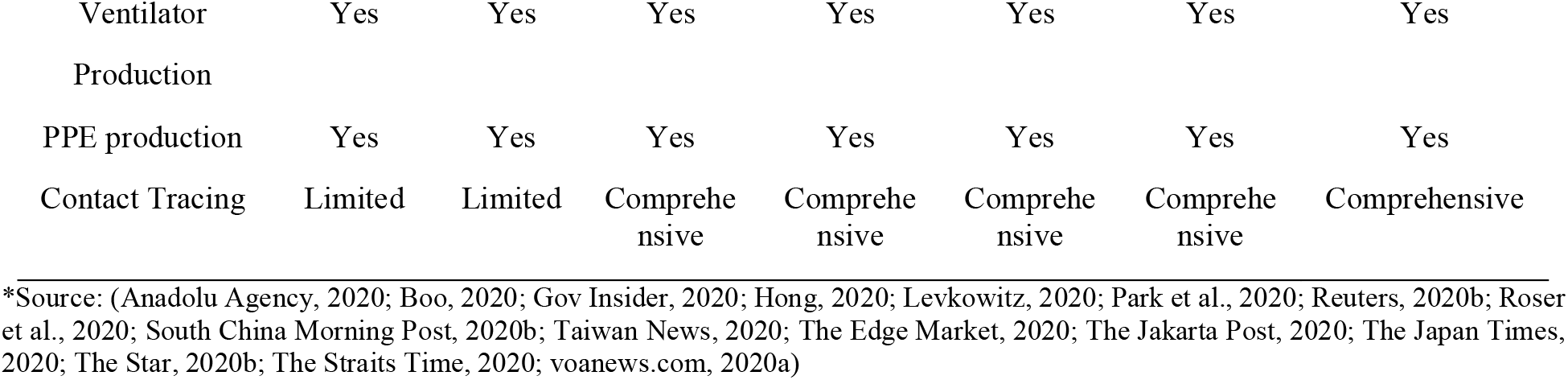
Technological availability for COVID-19 abatement.

Some countries had also developed its very own diagnostic kit that can detect COVID-19 to bolster testing rates in their respective country (Hong, 2020; The Star, 2020a; Woo, 2020). Indonesia’s kits could be mass-produced with a lower budget hence lowering expenditure (Taiwan News, 2020). Next, drive-thru testing is proven that this system is efficient as South Korea recorded a higher testing number as 50 drive-thru testing facilities were set up as of April 20 (Hong, 2020). In addition, most countries produce their own personal protective equipment (PPE) and ventilators as global supplies dwindle and most manufacturing companies focused on meeting their own country’s needs (Nikkei Asia, 2020). The amount of medical equipment should be proportional to the country needs to ensure all patients were given the chance to be treated and healed, thus decreasing mortality rate of the country.

### ECONOMY AND LIVELIHOOD

Based on Table 3, New Zealand, Singapore, South Korea, and Japan are classified as high-income countries. Meanwhile, Malaysia and Indonesia are categorized as upper middle-income and Vietnam as lower middle-income country. Having immense resource allows the government to invest in better facility, technology and better remuneration packages that covers broad relief during crisis as seen in Singapore (Woo, 2020). In addition, it allows the government to exercise stringent rules and have effective enforcement. To date, Indonesia has the highest percentage of poverty among the seven countries with half of its populations are struggling to make ends meet. A national lockdown may not be a preferred choice as it causes their economy to collapse, affecting their livelihood and putting tens of millions of its people who worked poorly paid informal jobs at risk (Aljazeera, 2020; New Straits Time, 2020).

### LOCAL LEADERSHIP PRESENCE

During the pandemic, the political system of each country plays a vital role in manoeuvring the nation through the pandemic. Table 8 further elaborating on Table 3 explains the local leadership status where P, PP, and A translates to absolute involvement, partial involvement, and least involvement, respectively. In Vietnam, the role of the government is very apparent as they were authoritarian in handling the pandemic. The government is more aggressive in enforcing laws without any delay and public debate (Cerre, 2020). The Vietnamese government had no problem mobilizing their military for health care missions while enforcing strict restrictions publicly. In an effort to ensure all citizens follow the social distancing and quarantine rule, Vietnam’s entrenched system of loyal neighbourhood party cadres had taken liberty to spy on residents and report their sightings to the superior (Jones, 2020). Kleinfield argues that the type of regime does affect a country’s success, but this alone does not sufficiently contribute to explaining the success (Kleinfeld, 2020).

**TABLE 8.** Local leadership attributes

|  | Indonesia | Japan | Malaysia | New Zealand | Singapore | South Korea | Vietnam |
| --- | --- | --- | --- | --- | --- | --- | --- |
| Democracy Index | Flawed Democracy | Flawed Democracy | Flawed Democracy | Full Democracy | Flawed Democracy | Flawed Democracy | Authoritarian |

\*Source: The Economist Intelligence Unit, 2020

### PENALTY

Table 9 outlines the compounds employed by the seven countries for potential offenses committed during the pandemic period. Most of the time, the amount of penalty is high to instil fear and compliance in the public. Thus, people are more inclined to adhere to the outlined SOPs consequently curbing the virus transmission risks. As opposed to other countries, Japan did not impose any penalty on its citizens as summarized in Table 3. Instead, it relies heavily on the people’s obedience (Bremmer, 2020; Nippon, 2020). The Japanese law had prohibited the government from enforcing stringent penalties as it was formed based on the idea that human rights should be respected. This can be linked with incidents during the World War Two where Japan had memories of civil rights abuse (Reuters, 2020a). Therefore, the government can only ‘request’ the people to minimize their movement. Social pressure is at work as the public averts risk of bearing responsibility for spreading the virus as they fear social sanctions (Nippon, 2020).

**TABLE 9.** A breakdown of penalties employed during COVID-19.

|  | Indonesia | Japan | Malaysia | New Zealand | Singapore | South Korea | Vietnam |
| --- | --- | --- | --- | --- | --- | --- | --- |
| Fleeing quarantine | 1-year / IDR 100 million | - | 2 - 5 years | \$4,000/6 months | SGD10,000/6 months, or both | ₩10 million /1-year (local), 3 year (foreigner) | VND 2,000,000 to VND 100,000,000 |
| Violating lockdown | 16-month | - | RM1000/6 | \$4,000/6 months | SGD10,000/6 months, or both | ₩3 million | VND 10,000,000 (individual) VND 20,000,000 (organization)/ VND 100,000,000 business closure |
| Failure complying to government personnel | - | - | RM10,000/2 years, or both | \$4,000/6 months | - | 5 years or maximum fine of ₩10 million | - |
| Food preparation. violation (absence mask/face shield) | - | - | - | - | \$5,000/suspension and license cancellation, or both | - | - |
| Spreading fake news | 6 years and maximum of IDR 1 million |  | RM2000-RM5000/5 years | - | Publish notices on false statements, provide link to government website with official clarifications | 5 years or maximum fine of ₩10 million | VND10-20 million |
| Mask | Warning | - | - | - | SGD300-SGD1000 | Corrective action | VND 300,000 and VND 7,000,000 if not discarded properly. |

\*Source: (Assegaf Hamzah & Partners, 2020; Astro Awani, 2020; Bremmer, 2020; Channel News Asia, 2020a; Dtinews.vn, 2020; Hong, 2020; Liew, 2020; Nasional Kompas, 2020; Nguyen & Pearson, 2020; Nippon.com, 2020; Reuters, 2020a; South China Morning Post, 2020a; Tang, 2020; The Edge, 2020)

### STRICT LOCKDOWN ENFORCEMENT

As cases spike, many countries resort to implementation of lockdown to contain the virus from spreading. Table 10 analysed that although most countries employed a strict national lockdown together with public area closures (P), some only endorsed closure of public areas and gathering bans (PP) while some practices none of them (A). Lockdowns become more effective especially when paired up with policies of social distancing and mandatory mask wearing. It proves to be a great circuit breaker as it restricts movements and proximity events among citizens. Therefore, possibility of a human transmission would be low, leading to lowering cumulative cases, daily case increase and death tolls.

**TABLE 10.**
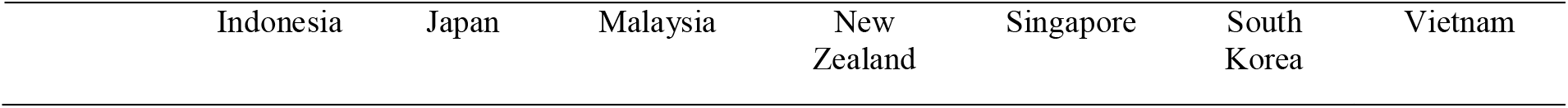
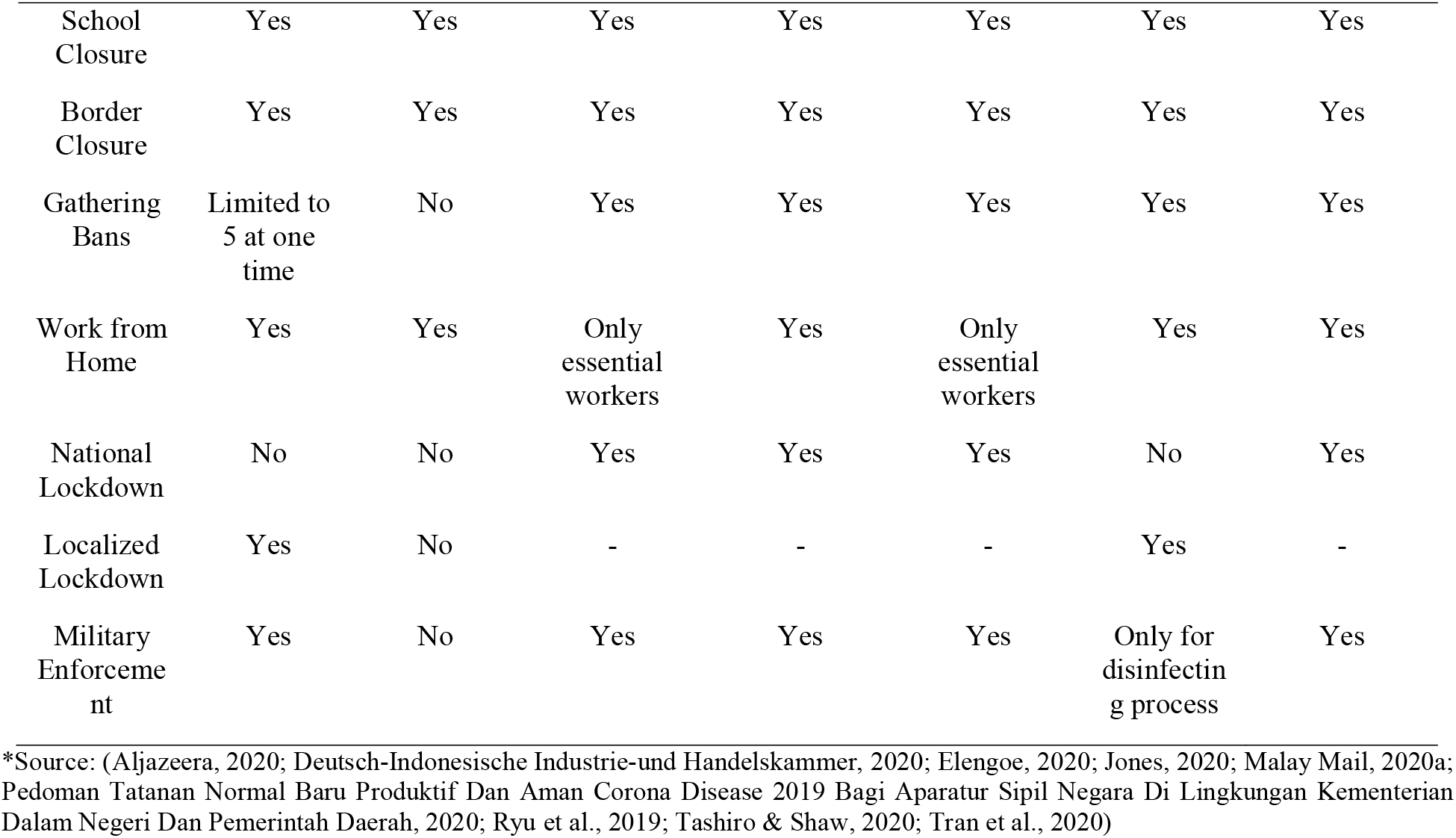
Lockdown enforcement criteria of each country.

| Indonesia | Japan | Malaysia | New Zealand | Singapore | South Korea | Vietnam |
| --- | --- | --- | --- | --- | --- | --- |

|  |  |  |  |  |  |  |  |
| --- | --- | --- | --- | --- | --- | --- | --- |
| School Closure | Yes | Yes | Yes | Yes | Yes | Yes | Yes |
| Border Closure | Yes | Yes | Yes | Yes | Yes | Yes | Yes |
| Gathering Bans | Limited to 5 at one time | No | Yes | Yes | Yes | Yes | Yes |
| Work from Home | Yes | Yes | Only essential workers | Yes | Only essential workers | Yes | Yes |
| National Lockdown | No | No | Yes | Yes | Yes | No | Yes |
| Localized Lockdown | Yes | No | - | - | - | Yes | - |
| Military Enforcement | Yes | No | Yes | Yes | Yes | Only for disinfecting process | Yes |

\*Source: (Aljazeera, 2020; Deutsch-Indonesische Industrie-und Handelskammer, 2020; Elengoe, 2020; Jones, 2020; Malay Mail, 2020a; Pedoman Tatahan Normal Baru Produktif Dan Aman Corona Disease 2019 Bagi Aparatur Sipil Negara Di Lingkungan Kementerian Dalam Negeri Dan Pemerintah Daerah, 2020; Ryu et al., 2019; Tashiro & Shaw, 2020; Tran et al., 2020)

In Japan, the government is incapable of enforcing lockdown due to the Japanese Law. However, an amendment has also been made to allow the Prime Minister to declare the ‘state of emergency’ in areas that are heavily inflicted by the virus. Both South Korea and Indonesia had applied a national social distancing policy, instead of a national lockdown (Table 10). Localized lockdowns will only be implemented at highly infected areas (Aljazeera, 2020; Ryu et al., 2019). However, the amount of comprehensive rapid testing done by South Korea had set their outcomes apart. Probably due to the poorer state of the economy, Indonesia’s response was slow and unclear, which in the end had costed them a spike in cases.

### STRICT STANDARD OF PROCEDURE IN PUBLIC AREAS

Table 3 shows that all countries were coded P, with implementation of all SOPs except for the mask wearing requirement (see Table 11). SARS-CoV-2 spread by water droplets which could be suspended in air or on surfaces. The WHO had suggested mask wearing as an integral strategy to suppress transmission coupled with other policies such as social distancing, health checks and temperature scanning in public areas, washing hands and sterilization. Sterilization and fumigation procedures are vital at frequently used public spaces, such as public transportation stations, shops, and especially infected venues. These SOPs are crucial as they can break the virus chain if abided accordingly, hence lowering infection cases.

**TABLE 11.**
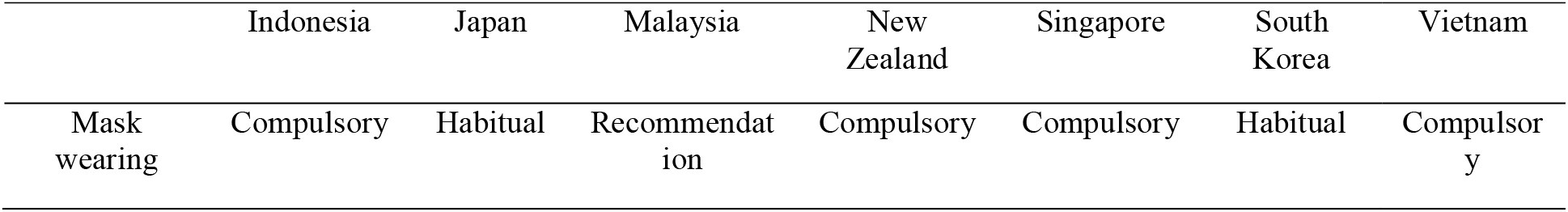
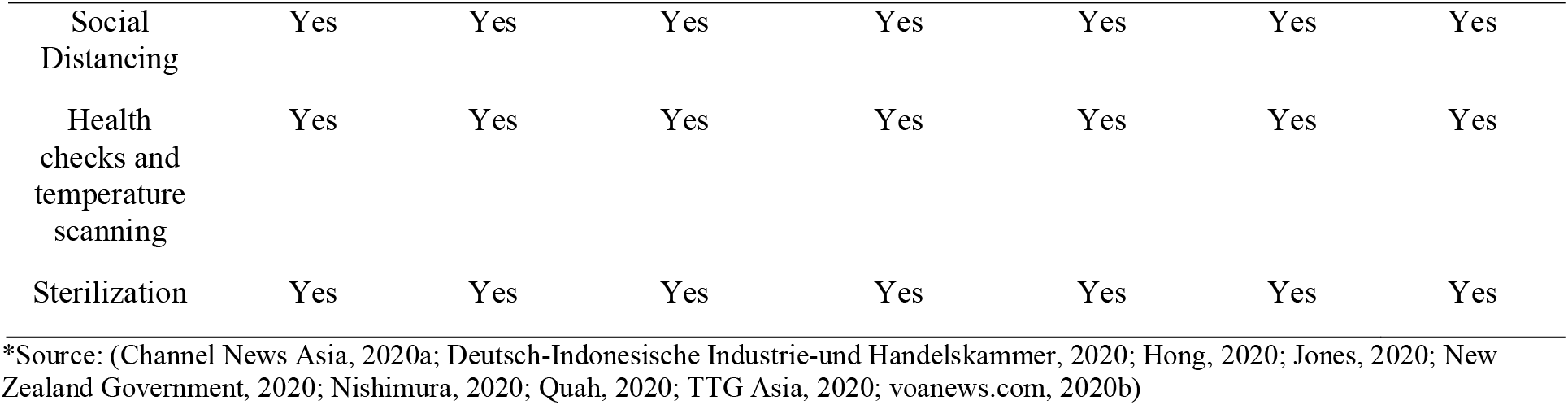
SOPs in respective countries.

### EMERGENCY RESPONSE PLANS

An emergency response plan in a pandemic situation is crucial to ensure an effective control of the pandemic. Table 3 shows that all countries had devised their own emergency response plan, denoted as P. Among popular forms of emergency response plan are debt relief and income support by the government as stated in Table 12. Having a broad relief helps decrease the citizen’s movement as they can stay home without worrying much about their financial conditions. Restricting movement proves to be an excellent circuit breaker to the virus transmission.

**TABLE 12.**
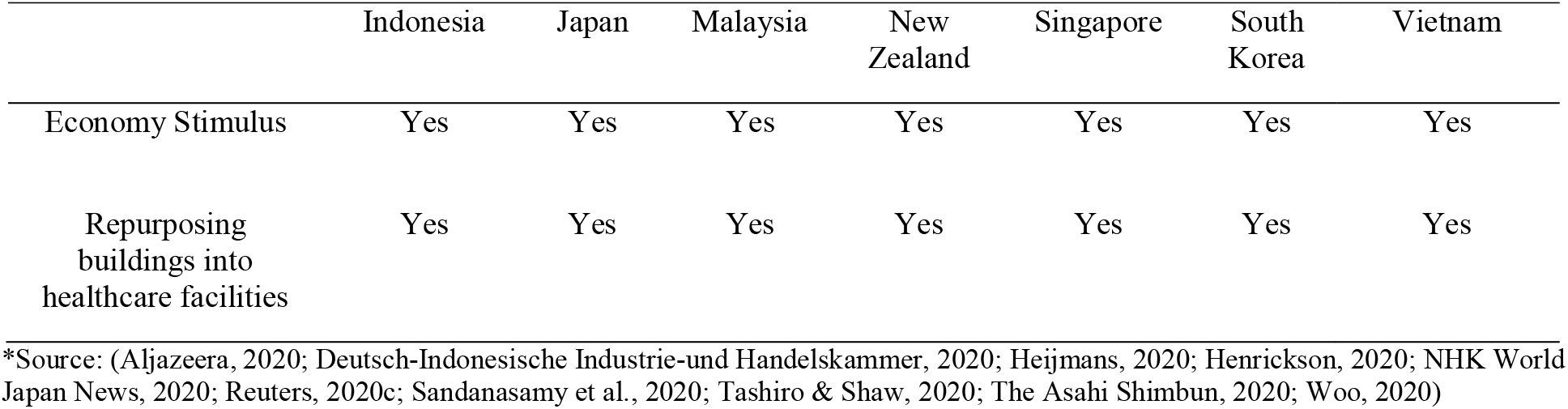
Countries’ emergency response plans.

As a step to preparing for the worse, the governments also called up for the increase of healthcare facilities by turning buildings into quarantine centres, mobilizing more labs for testing and research purposes (Abe, 2020; Ngoc Thanh & Tat Dinh, 2020; Rahman, 2020). For example, Indonesia had repurposed a truck into a laboratory to increase testing rates (The Jakarta Post, 2020). The Korean and Malaysian healthcare had also transformed a few hospitals into special COVID-19 hospitals (Elengoe, 2020; Her, 2020); thus, specialized healthcare workers could prioritize treating COVID-19 patients until recovery.

### INTERACTION ARENA

In addition to the SES attributes described above, efficient monitoring, communication, movement restriction and extensive contact tracing and testing as part of the interaction arena (activities/process as a result of the SES attributes), sourced from the Government Stringency Index by the Oxford COVID-19 Government Response Tracker, were used as supplemental evidence and reference to further explain the outcome of each country. The same coding system shown in the analysis section was used, where code P was used to represent data with 70-100 score. Meanwhile, PP represents data with 40-69 score, while A denotes a score ranging 0-39. The data varied every month; hence, the highest score was considered when assigning the code. New Zealand had recorded the highest stringency score at 96, followed by Singapore at 85, Vietnam at 83, Indonesia at 80, South Korea and Malaysia at 75, and Japan at 47. A high stringency reflects an effective monitoring, in general. Meanwhile, Singapore had the highest containment index with a score of 86, followed by New Zealand at 83, South Korea at 79, Malaysia at 77, Indonesia at 73 and finally Japan at 44. Furthermore, Singapore had the highest testing rate, followed by New Zealand, Vietnam, Malaysia, South Korea, Japan and lastly Indonesia. Also, all countries practice a coordinated information campaign, which promotes effective communication between the government and citizens.

## CONCLUSION AND RECOMMENDATIONS

The primary objective of this study was to explain COVID-19 from an SES perspective, hence testing out the framework’s adaptability in the health and medical setting. Through the framework, seven Asia Pacific countries were reviewed for institutional-socio-ecological factors which affect their success in fighting the pandemic. Due to the adherence to or high presence (P) frequency of the design principles, both Vietnam and New Zealand classified as countries with the high success level of COVID-19 abatement are coherently explainable. Meanwhile, Japan and Indonesia, with low presence (P) frequency, were deemed as countries with the low success level.

Although some countries possessed a high number of Ps for certain design principles/attributes, the outcome signified that they may not necessarily and ideally abate the pandemic (e.g., South Korea, Singapore, and Malaysia) as what have been hypothesised. Thus, three lessons can be drawn: (i) having high number/frequency of Ps for SES attributes does not always constitute them a panacea for the pandemic; (ii) however, although the presence of attributes may not always lead to success, it would be detrimental to a country for not having them; and lastly (iii) some attributes (mostly from the governance factor) may have a heavier weightage (more significant) towards explaining the success level. More precisely, the common effective traits (or critical success factors/design principles) found in Vietnam and New Zealand are ultimately about the implementation of lockdown, penalties, strict public standard of procedures, strong leadership, trust and reciprocity between government and citizens, comprehensive testing, efficient mobilized facilities, and past experiences in other related contagions. The economic situation of a country does play a vital role in keeping the pandemic under control but Vietnam demonstrated otherwise or is an ‘anomaly’ in handling their COVID-19 situation, given its status as a lower middle income nation by the World Bank. Meanwhile, low success outcome countries such as Japan and Indonesia are associated with the lack of national lockdown implementation and low testing rates, and specifically in Indonesia, due to its poverty and economic situations, it faced inadequate facilities and resources, and manpower to contain the pandemic. Other factors such population size and density, effective foreign workers’ influx containment, homogeneity, and emergency response plan are not as impactful and significant as the above factors.

By understanding and identifying which are the most significant SES factors, this comparative study hopes to shed light on how to leverage on those institutional-social-ecological attributes so that more strategic economic policy can be devised by policymakers address the crisis. The SES framework indeed acts as a common language among researchers, hence bridging boundaries of different disciplines. However, this research was primarily limited to literature reviews and secondary data analysis, subject to certain level of uncertainties, subjectivity and data inconsistency and unavailability. Future studies should focus on the inclusion and further verification of the existing and other SES attributes (higher-tier SES) to explain and predict the pandemic outcome, as well as the adaptation of the IAD-SES framework into other health-related topics.

## Data Availability

All data are available on the internet which have been listed out in the reference section

## ACKNOWLEDGMENT

The authors would like to express utmost gratitude to Universiti Teknologi Malaysia, Johor Bahru for their support and cooperation.

## Footnotes

1 SES and IAD-SES will be used interchangeably throughout the paper.

## Notes

### Competing Interest Statement

The authors have declared no competing interest.

### Author Declarations

Exemption has been granted

